# Mortality is lowest in overweight followed by obese and morbid obese patients and is highest in cachexia compared to normal weight in patients with the diagnosis of aortic stenosis

**DOI:** 10.1101/2024.08.17.24312157

**Authors:** Mehrtash Hashemzadeh, Arman Soltani Moghadam, Saman Soltani Moghadam, Mohammad Reza Movahed

## Abstract

**Introduction:** The obesity paradox has been seen in many cardiovascular conditions. The goal of this study was to evaluate whether it exists in patients with a diagnosis of aortic stenosis.

**Method:** We used the Nationwide Inpatients Sample (NIS) database and ICD-10 coding for adults in different weight categories and with aortic stenosis diagnoses for 2016-2020. We evaluated the effect of weight on mortality using multivariate adjustment and the cox-regression model.

**Results:** A total of 2,330,584 patients were diagnosed with aortic stenosis. Mortality was lowest in overweight followed by obesity and morbid obesity (1.74% vs. 2.43% vs 3.2% in comparison to normal weight mortality of 4.4%, p<0.001) and it was highest in patients with cachexia (mortality of 14.5%). After adjusting for baseline characteristics and comorbid conditions, the relation between mortality and weights remained unaltered. Multivariate adjusted odds ratios (OR) were as follows: Overweight OR 0.4, CI 0.31-0.6, p<0.001, Obesity: OR 0.64, CI 06-0.68, p<0.001, morbid obesity OR: 0.88, CI 0.83-0.94, P<0.001, Cachexia OR 3.31 CI: 3.04-3.62, p<0.001).

**Conclusion:** Using the largest database, we found that in patients with a diagnosis of aortic stenosis, overweight followed by obesity and morbid obesity have the lowest mortality whereas cachexia has the highest mortality compared to normal-weight patients.

## Introduction

Contributing to increased mortality and morbidity, obesity has proved to be a well-established risk factor for many diseases(1). The incidence of obesity, typically measured by body mass index (BMI), has been rising sharply(2). According to current data, over two-thirds of adults in the United States and more than 2.1 billion individuals globally are affected by obesity (2, 3). The ’obesity paradox’ describes a situation where, surprisingly, obesity appears to be beneficial and linked to improved survival rates in some patient groups. This phenomenon has been consistently observed in patients undergoing percutaneous coronary intervention (PCI) or coronary artery bypass grafting (CABG) for ischemic heart disease(4, 5). Recently, the obesity paradox has also been observed in other types of cardiac surgeries and interventions, such as surgical and transcatheter aortic valve replacement (TAVR) for treating severe aortic stenosis (AS) (6–8). AS is the most prevalent valvular heart disease in Western developed countries. As Western populations age, the negative impacts on clinical symptoms, mortality, and economic costs are anticipated to rise (9, 10). Notably, a significant number of obese patients with severe AS are scheduled for TAVR, and this number is expected to grow as the population ages(11).

In patients with a diagnosis of AS, controversy exists regarding the effects of body weight on mortality, and the exact effects remain elusive(12–14). Using the Nationwide Inpatient Sample (NIS) database, we sought to evaluate the impact of body weight on mortality in patients with AS and show whether the obesity paradox exists in these patients.

## Methods

### Data Source

This study utilized the NIS database, part of the Healthcare Cost and Utilization Project (HCUP) in the United States. The NIS is a publicly available resource extensively used by researchers and policymakers. It encompasses approximately 20% of all hospital discharges for adults in the United States, featuring data from around 7 million unweighted and 35 million weighted hospital stays annually. The NIS analyzes healthcare trends, utilization, costs, and outcomes. The dataset is de-identified and exempt from Institutional Review Board (IRB) approval. It includes comprehensive information on primary and secondary diagnoses, procedures, discharge status, and demographic details from a representative sample of community hospitals.

### Sample Selection

This retrospective study included adults aged 18 and older hospitalized with an AS diagnosis between 2016 and 2020. The study population was identified using specific International Classification of Diseases, Tenth Revision (ICD-10) codes for aortic stenosis. Patients were categorized based on different weight classes as determined by ICD-10 coding for obesity (E66.9, E66.8, E66.0), overweight (E66.3), morbid obesity (E66.01, E66.2), and cachexia (R64). Body weight categories were recorded from a chart based on BMI extracted from the chart used by coders consistent with different weight categories. The primary outcome evaluated was all-cause mortality. To mitigate the impact of confounding variables, a multivariate analysis was conducted, adjusting for different variables including age, sex, smoking status, diabetes, hypertension, history of myocardial infarction, chronic obstructive pulmonary disease (COPD), chronic kidney disease (CKD), aortic valve surgery, and race.

### Statistical Analysis

Patient demographic, clinical, and hospital characteristics were reported as median (IQR) for continuous variables and proportions, with 95% confidence intervals for categorical variables. Logistic regression was performed to ascertain the odds of binary clinical outcomes relative to patient and hospital characteristics as well as ascertaining the odds of clinical outcomes over time. All statistical models were adjusted for confounding. All analyses were conducted following the implementation of population discharge weights. All p-values were 2-sided and p<0.05 was considered statistically significant. Data were analyzed using STATA 17 (Stata Corporation, College Station, TX).

## Results

A total of 2,330,584 patients (51% male) with the diagnosis of AS were included in this study. The median age of the population was 80 years (IQR=72-87) and 71.49% of the patients were visited in urban teaching hospitals. The complete baseline characteristics of the patients are available in **Table 1**. The overall mortality rate was estimated to be 4.20% of the total population. Compared with normal body weight, the mortality was lower in overweight individuals (1.74% vs 4.40%, OR=0.38, 95%CI 0.28-0.53, p<0.001). Similarly, the mortality was lower in patients with obesity (2.43% vs 4.40%, OR=0.54, 95%CI 0.51-0.57, p<0.001) and morbid obesity (3.20% vs 4.40%, OR=0.72, 95%CI 0.68-0.76, p<0.001). Conversely, patients with cachexia had a statistically significant higher mortality rate in comparison with those with a normal body weight (14.56% vs 4.40%, OR=3.70, 95%CI 3.20-4.02, p<0.001) (**Table 2**). The time trend in mortality within each group from 2016 to 2020 is shown in **Figure 1-4**.

**Figure 1:**
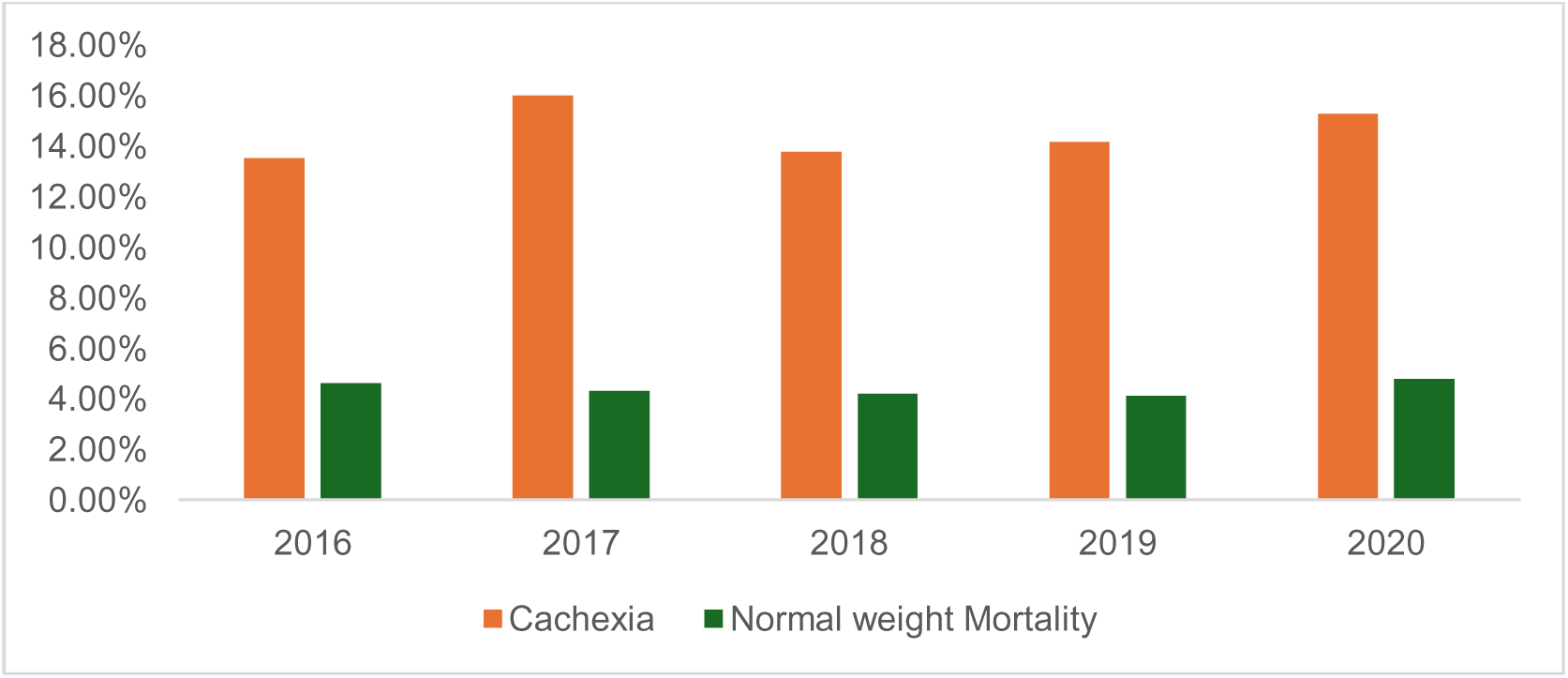
Time trends in mortality from 2016-2020 based on the cachexia vs normal weight

**Figure 2:**
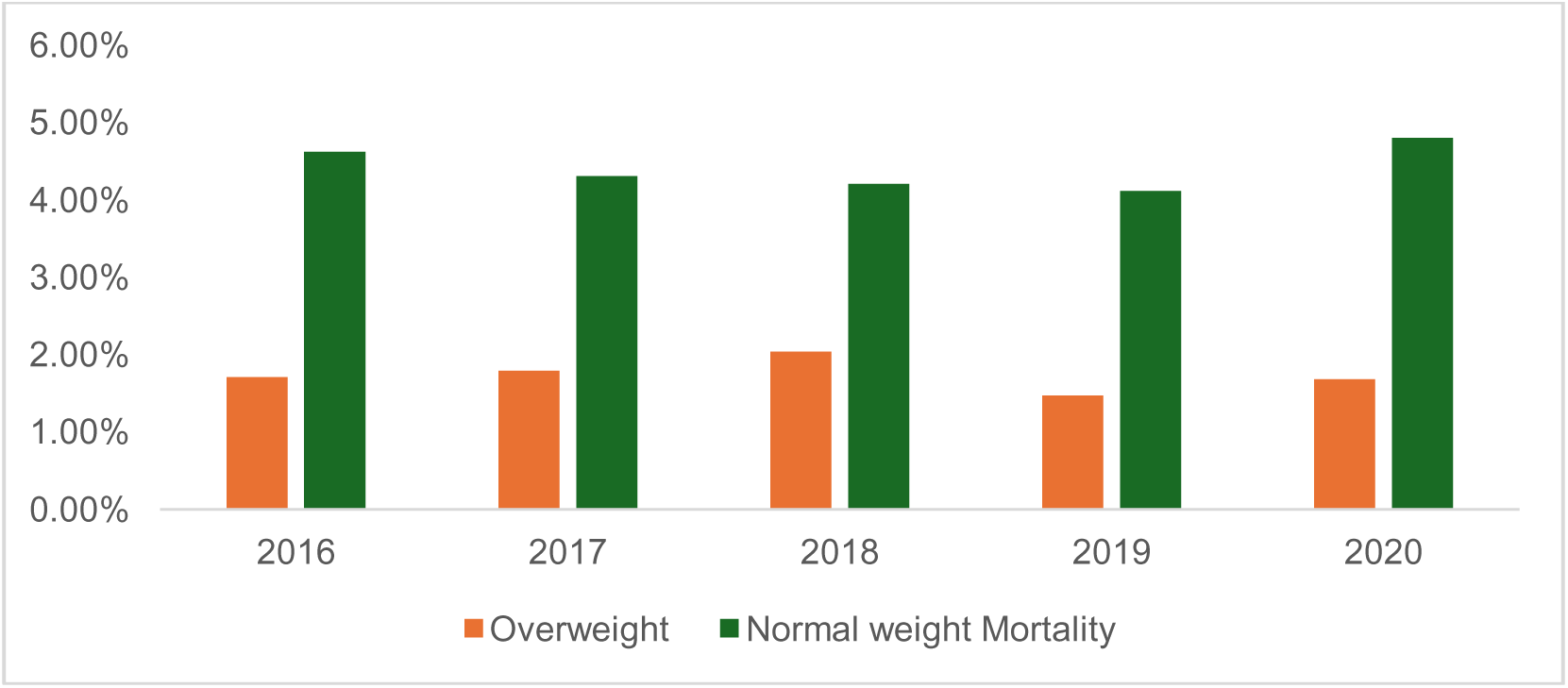
Time trends in mortality from 2016-2020 based on the overweight vs normal weight

**Figure 3:**
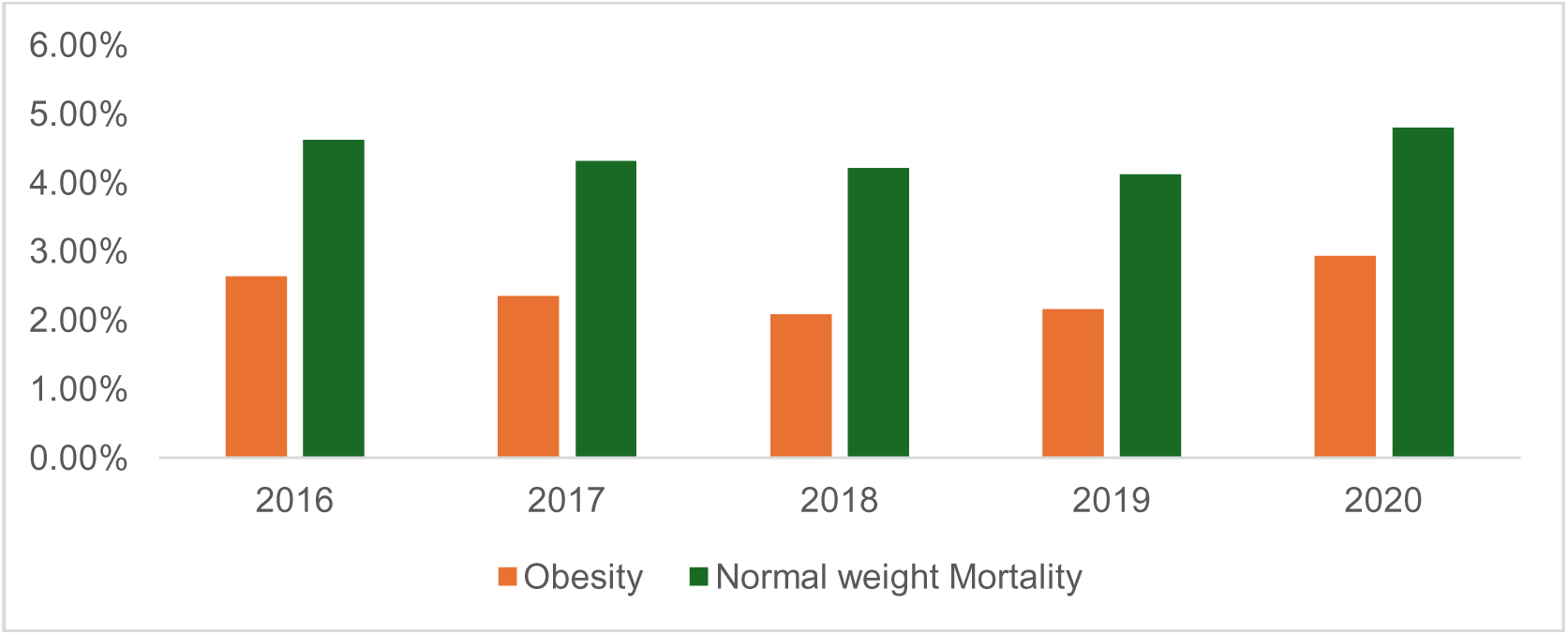
Time trends in mortality from 2016-2020 based on the obesity vs normal weight

**Figure 4:**
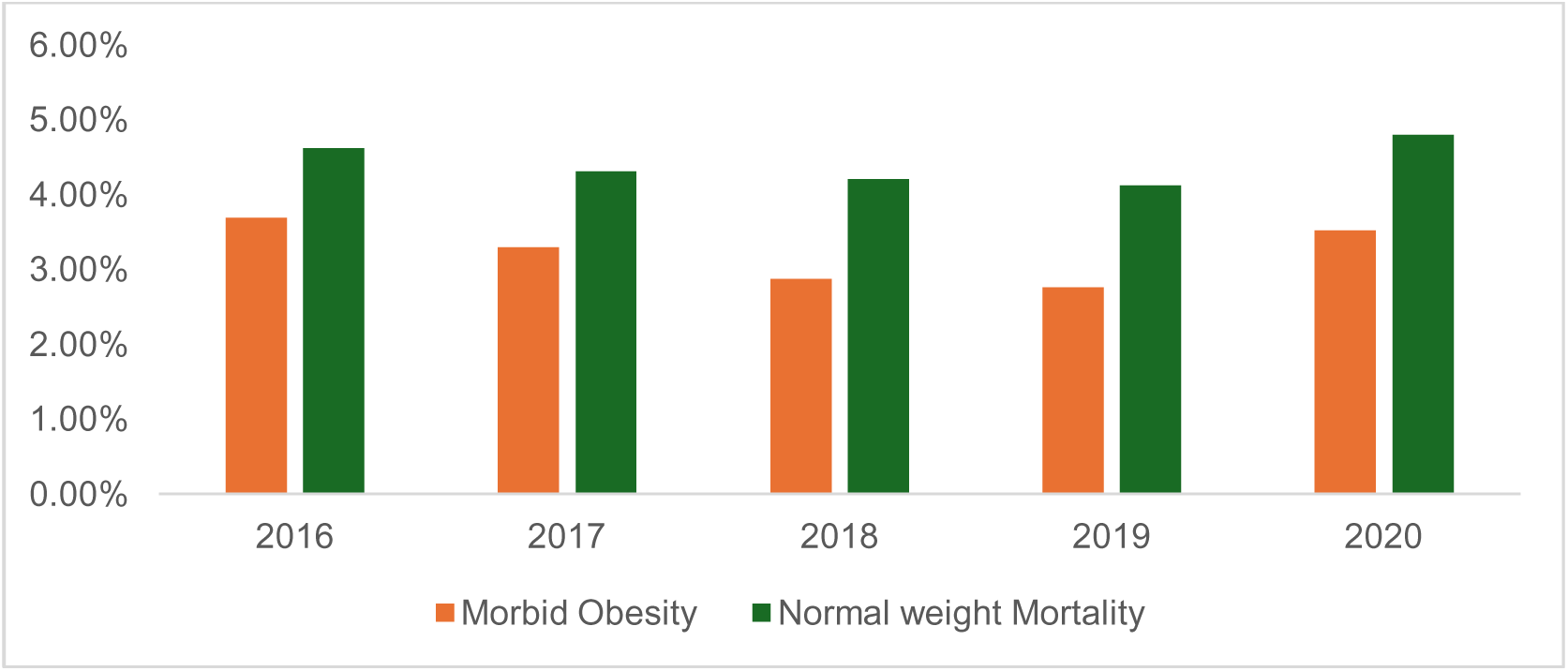
Time trends in mortality from 2016-2020 based on the morbid obesity vs normal weight

**Figure 5:**
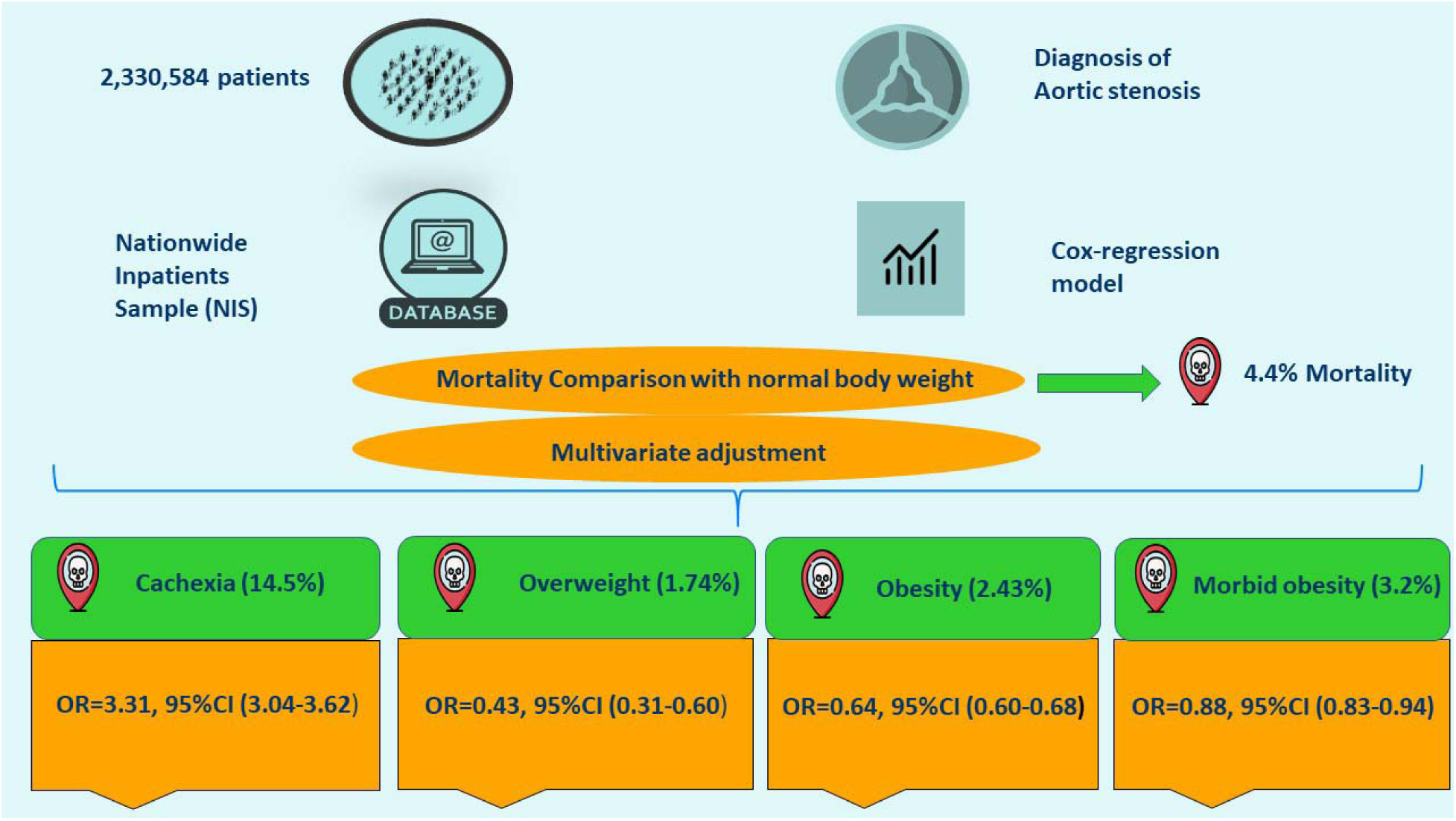
Comparing mortality data of all weight categories

**Table 1:** Basline Charcteristics.

| AGE above 18 |  |  |  |  |  |  |
| --- | --- | --- | --- | --- | --- | --- |
| 2016-2020 | Total Aortic Stenosis | Normal Weight | Cachexia | Overweight | Obesity | Morbid Obesity |
| Total Population | 2,330,584 | 1,885,709 | 23,535 | 11,505 | 230,195 | 182,005 |
| Age (Median(IQR)) | 80 (72-87) | 82 (74-86) | 85 (76-90) | 78 (71-85) | 75 (68-82) | 72 (65-78) |
| LOS (Median(IQR)) | 4 (2-7) | 4 (2-7) | 5 (3-9) | 4 (2-7) | 4 (2-7) | 5 (3-8) |
| Total Charges \$ (Median(IQR)) | 50385 (25245-111984) | 48397.5 (24415-109295) | 48687 (25098-73643) | 61052 (29136-142295) | 62367 (29947-136156) | 58572 (29306-128738) |
| Mortality | 1.20% | 1.10% | 11.56% | 1.71% | 2.13% | 3.20% |
| Gender |  |  |  |  |  |  |
| Male | 51.24% | 52.21% | 48.03% | 57.00% | 51.43% | 41.21% |
| Female | 48.74% | 47.79% | 51.97% | 43.00% | 48.57% | 58.79% |
| Race |  |  |  |  |  |  |
| White | 83.34% | 83.32% | 80.01% | 83.68% | 84.05% | 83.09% |
| Black | 5.53% | 6.32% | 8.36% | 5.33% | 6.84% | 8.18% |
| Hispanic | 5.05% | 6.04% | 5.80% | 6.61% | 6.15% | 6.07% |
| Asian Pac Isl | 1.66% | 1.85% | 3.15% | 1.78% | 0.78% | 0.48% |
| Native American | 0.32% | 0.31% | 0.30% | 0.09% | 0.38% | 0.41% |
| Other | 2.09% | 2.16% | 2.38% | 2.21% | 1.80% | 1.67% |
| Smoking | 31.74% | 31.80% | 26.36% | 38.67% | 35.34% | 31.45% |
| Diabetes | 36.42% | 36.59% | 22.43% | 42.50% | 56.50% | 65.65% |
| Hypertension | 86.62% | 86.71% | 79.39% | 89.74% | 91.84% | 91.48% |
| COPD | 25.03% | 24.21% | 35.90% | 23.16% | 27.66% | 36.26% |
| CKD | 38.23% | 38.23% | 37.92% | 36.55% | 39.14% | 43.30% |
| STEMI | 3.10% | 3.08% | 4.31% | 2.52% | 2.85% | 2.73% |
| NON-STEMI | 7.20% | 7.20% | 6.95% | 7.48% | 7.01% | 6.21% |
| CADMI | 11.51% | 11.52% | 10.78% | 11.73% | 12.47% | 10.11% |
| Aortic Valve Surgery | 5.03% | 5.09% | 0.62% | 10.51% | 10.16% | 7.31% |

**Table 2:**
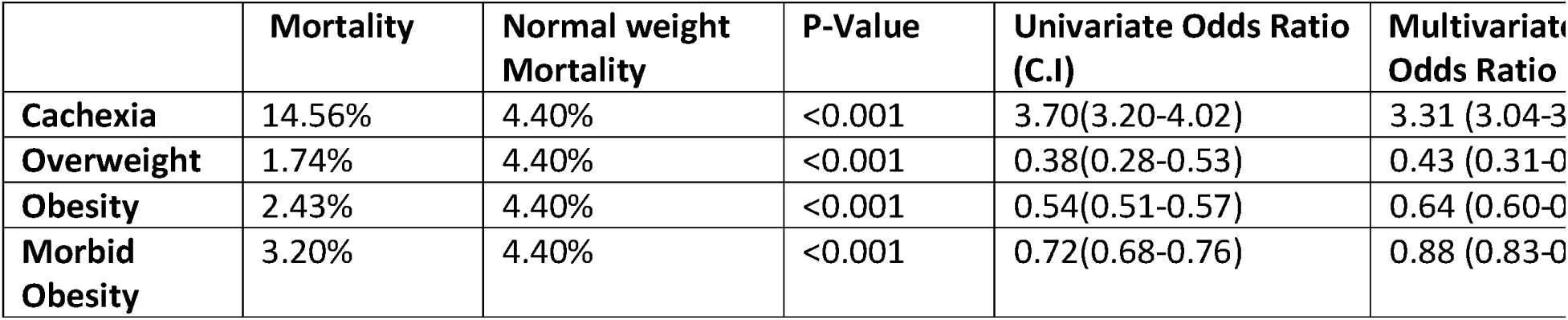
Univariate and Multivariate analysis for the cox-regression model comparing mortality with normal-weight patients:

|  | Mortality | Normal weight Mortality | P-Value | Univariate Odds Ratio (C.I) | Multivariate Odds Ratio |
| --- | --- | --- | --- | --- | --- |
| Cachexia | 14.56% | 4.40% | <0.001 | 3.70(3.20-4.02) | 3.31 (3.04-3 |
| Overweight | 1.74% | 4.40% | <0.001 | 0.38(0.28-0.53) | 0.43 (0.31-0 |
| Obesity | 2.43% | 4.40% | <0.001 | 0.54(0.51-0.57) | 0.64 (0.60-0 |
| Morbid Obesity | 3.20% | 4.40% | <0.001 | 0.72(0.68-0.76) | 0.88 (0.83-0 |

After performing a multivariate analysis, the results of our study stayed consistent, and overweight patients demonstrated the lowest mortality (OR=0.43, 95%CI 0.31-0.60, p<0.001), followed by obese (OR=0.64, 95%CI 0.60-0.68, p<0.001) and morbid -obese patients (OR=0.88, 95%CI 0.83-0.94, p<0.001) compared to normal-weight patients. Notably, the mortality was highest in those with cachexia (OR=3.31, 95%CI 3.04-3.62). (**Table 2**)

## Discussion

Our study, which utilized the largest population sample to date from the NIS database, reveals significant findings on the paradoxical effects of body weight on mortality in patients with AS. This comprehensive analysis, the largest of its kind, demonstrated that body weight exerts a notable influence on mortality outcomes, even after adjusting for multiple variables. Specifically, cachexia was associated with the highest mortality risk compared to normal body weight (OR=3.31, 95% CI: 3.04-3.62). Conversely, being overweight was linked to the lowest mortality risk (OR=0.43, 95% CI: 0.31-0.60), followed by obesity (OR=0.64, 95% CI: 0.60-0.68) and morbid obesity (OR=0.88, 95% CI: 0.83-0.94). These findings underscore the complex relationship between body weight and survival outcomes in AS patients.

The paradoxical effects of body weight on mortality in patients with various conditions have been established in several studies. In patients undergoing revascularization strategies like PCI and CABG, obese patients tended to demonstrate better outcomes compared with those with a normal body weight(15–19). Furthermore, recent studies have proven that the paradox of obesity exists in the realm of TAVR with a meta-analysis suggesting a strong association between obesity and decreased mortality and cachexia with increased mortality (20, 21). This could be justified by several factors. First, particular challenges like worse vascular access due to the smaller size of the vessels could be encountered in patients with lower body weight which could further increase vascular complications and mortality(22). Secondly, in surgical patients, adipose tissue is thought to play a pivotal role in producing receptors of tumor necrosis alpha factors, thereby reducing the associated inflammation and providing better recovery and wound healing(23, 24).

However, previous studies have demonstrated conflicting results regarding the effects of body weight on mortality outcomes in patients with AS. A study by Rogge et.al (13) evaluating the effects of obesity on mortality in 1,664 patients with asymptomatic AS (SEAS study), demonstrated conflicting results with our study. They found that overweight and obese patients had higher rates of hypertension and abnormal left ventricular geometry but did not show a difference in AS progression rate between BMI classes. Univariate analysis suggested overweight was associated with a lower rate of AS-related and ischemic cardiovascular events. However, multivariate analyses indicated that being overweight had no significant impact on these events and was associated with higher rates of total mortality and combined heart failure hospital stays and death. This study denied the BMI paradox in AS, but it had notable limitations. The population was asymptomatic and relatively young, limiting the generalizability of their findings. Additionally, the SEAS study excluded patients with cardiovascular disease, diabetes, reduced LVEF, renal impairment, or other major valvular diseases, making the results inapplicable to these subpopulations. These limitations, along with conflicting results based on univariate versus multivariate analyses, highlight the complexity of assessing the impact of body weight on AS outcomes. On the other hand, similar to our findings, in a study by Ngiam et al. (12), 154 patients with medically managed severe AS were evaluated for the effects of body weight on mortality. After multivariate adjustment, obese patients remained protected against mortality (HR=0.38, 95%CI 0.15-0.98, p=0.04). Nonetheless, their study was mostly limited by the relatively small sample size and a heterogeneous Asian population. Moreover, they only divided patients into obese and non-obese. Unlike this study, our study comprised the largest sample size of 2,330,584 patients and we comprehensively assessed the effects of body weight in 5 different categories (normal weight, overweight, obese, morbidly obese, and cachexic) and compared them with normal-weight patients. Rossi et.al (14) conducted a study evaluating the effects of body weight on mortality in AS patients, and their results go in line with ours. They showed that decreased BMI was strongly associated with the highest mortality (HR 4.5, 95% CI 1.7–11.8; p = 0.002) compared to normal body weight.

The exact mechanism behind these mysterious paradoxical effects of body weight on mortality in patients with AS is yet to be explicitly understood. Nutritional depletion is thought to be the cornerstone of worsened clinical outcomes in cachectic patients. Although frailty and cachexia have been considered risk factors for worsening clinical outcomes in cardiology, decreased body weight could also be an inevitable consequence of a vicious cycle triggered by the progression of severe AS to advanced heart failure and cardiac cachexia(25–27).

Several factors may explain the decreased mortality observed in patients with higher body weights. Firstly, the increasing prevalence of obesity at a younger age, driven by the popularity of fast food and reduced physical activity, means that obese patients tend to be younger. Younger individuals generally have stronger physiological functions and better recovery capabilities, allowing them to tolerate surgical interventions better (28). Secondly, obese individuals are often treated more aggressively for their comorbid conditions related to obesity. Physicians frequently advise these patients to engage in regular exercise and follow a healthy diet, which is less common advice for normal-weight patients. Consequently, obese patients might be more vigilant about their health, seeking medical care sooner and thus obtaining earlier diagnoses for their conditions(17). Furthermore, the higher prevalence of cardiovascular risk factors such as hypertension, hyperlipidemia, and diabetes mellitus in obese patients leads to more intensive treatment with cardioprotective medications. These patients are also more likely to have closer follow-ups with a cardiologist and adhere to guideline-based medical therapy, which could contribute to better management of their health conditions and potentially confer a survival benefit(17, 29). Lastly, patients with higher BMI may be earlier in the course of their disease, whereas those with more advanced disease may exhibit symptoms of heart failure and cardiac cachexia, resulting in a lower BMI. This could create an apparent protective effect of higher BMI in patients with severe AS, rather than obesity exerting a true survival benefit.

These factors collectively suggest that the observed lower mortality in patients with higher body weights might be due to a combination of younger age, proactive healthcare management, and closer medical follow-up, rather than a direct survival benefit conferred by obesity itself.

Future research should focus on prospective studies to better elucidate the causal relationships between body weight and mortality in aortic stenosis patients. Additionally, exploring the underlying mechanisms and potential interventions to optimize outcomes for different weight categories will be crucial in advancing clinical practice

## Limitations

Our study has several limitations that should be acknowledged. First, the NIS database relies on the accuracy and completeness of ICD-10 coding, which may introduce potential misclassification or coding errors. Second, the observational and retrospective nature of the study precludes the establishment of causal relationships between body weight and mortality outcomes Furthermore, residual confounding factors might still influence the results despite adjusting for multiple confounders. Lastly, the study population primarily consisted of older adults hospitalized with AS, which may limit the generalizability of the findings to younger or community-dwelling populations.

## Conclusion

Our study confirms that the obesity paradox exists in patients with a diagnosis of AS. Cachectic patients carry the most risk and are associated with the highest mortality rate. Notably, overweight patients demonstrated the lowest mortality rate compared to patients with a normal body weight, followed by obesity and morbid obesity.

## Data Availability

NIS data publically available

